# Dynamic connectivity predicts acute motor impairment and recovery post-stroke

**DOI:** 10.1101/2020.09.25.20200881

**Authors:** Anna K. Bonkhoff, Anne K. Rehme, Lukas Hensel, Caroline Tscherpel, Lukas J. Volz, Flor A. Espinoza, Harshvardhan Gazula, Victor M. Vergara, Gereon R. Fink, Vince D. Calhoun, Natalia S. Rost, Christian Grefkes

## Abstract

**Objective:** Thorough assessment of cerebral dysfunction after acute brain lesions is paramount to optimize predicting short- and long-term clinical outcomes. The potential of dynamic resting-state connectivity for prognosticating *motor* recovery has not been explored so far.

**Methods:** We built random forest classifier-based prediction models of acute upper limb motor impairment and recovery after stroke. Predictions were based on structural and resting-state fMRI data from 54 ischemic stroke patients scanned within the first days of symptom onset. Functional connectivity was estimated using both a static and dynamic approach. Individual motor performance was phenotyped in the acute phase and six months later.

**Results:** A model based on the time spent in specific dynamic connectivity configurations achieved the best discrimination between patients with and without motor impairments (out-of-sample area under the curve and 95%-confidence interval (AUC±95%-CI): 0.67±0.01). In contrast, patients with moderate-to-severe impairments could be differentiated from patients with mild deficits using a model based on the variability of dynamic connectivity (AUC±95%-CI: 0.83±0.01). Here, the variability of the connectivity between ipsilesional sensorimotor cortex and putamen discriminated the most between patients. Finally, motor recovery was best predicted by the time spent in specific connectivity configurations (AUC±95%-CI: 0.89±0.01) in combination with the initial motor impairment. Here, better recovery was linked to a shorter time spent in a functionally integrated network configuration in the acute phase post-stroke.

**Interpretation:** Dynamic connectivity-derived parameters constitute potent predictors of acute motor impairment and post-stroke recovery, which in the future might inform personalized therapy regimens to promote recovery from acute stroke.

## Introduction

Stroke is the leading cause of long-term disability in adults^1^ and entails the highest number of disability-adjusted life years among more than 300 different causes.^2^ To optimize stroke care, it is of great importance to establish prediction models of stroke-related disabilities at the single patient level. These predictions may not only inform patients and their proxies about individual trajectories after stroke, but may also facilitate the allocation and planning of targeted rehabilitative regimens.

The amount of initial motor impairment has been frequently demonstrated to constitute a strong predictor of chronic impairment,^3^ yet may not guarantee sufficiently accurate predictions at the level of single patients.^4^ This limitation motivates the consideration of further biomarkers of stroke recovery.^5^ Structural and functional neuroimaging-derived information have proven viable candidates: For example, we have demonstrated that recovery after stroke could be reliably predicted by functional neuroimaging signals obtained during a motor task.^6^ Task-based data, however, depends critically on the patient’s ability to accurately perform a task of interest. Given this requirement, patients with severe impairments are frequently excluded a-priori from such studies. In contrast, structural and resting-state functional magnetic resonance imaging (rsfMRI) data depend much less on the abilities of a patient. Since both modalities do not require active participation or specific stimulation equipment, they may be easily implemented into the clinical routine, also allowing to consider patients with a wide range of clinical deficits. While previous studies relying on either structural^7^ or resting-state MRI data^8^ have generated promising results for the prediction of motor impairments early after stroke, data are scarce with respect to successfully predicting motor recovery as a long-term outcome. One reason might lie in the relatively low sensitivity and temporal resolution of conventional rsfMRI analyses assuming static connectivity between brain areas. However, novel techniques permit the increase in the temporal resolution of resting-state connectivity maps from minutes to seconds.^9,10^ By these means, dynamic functional network connectivity (dFNC) analyses are thought to capture moment-to-moment fluctuations in a behaviorally meaningful way^11,12^ and grant novel insights into neurological disease (e.g., epilepsy,^13^ migraine,^14^ Parkinson’s disease,^15^ Huntington’s disease^16^). We have recently shown that dFNC analyses provide new information on cerebral alterations post-stroke, thus far hidden in static analyses.^17^ Several studies suggest that dFNC may not only enhance inferences in imaging correlates of brain disease but also lead to substantial increases in predictive capacities for several of these diseases, potentially as the dynamic approach may to be closer to the neurobiology underlying brain function.^18,19,20^

Therefore, we here sought to explore the potential of dFNC parameters for predicting acute motor impairment and recovery after stroke. We hypothesized that dFNC parameters obtained in the acute post-stroke phase are superior to static connectivity for predicting motor outcome as neural reorganization enabling recovery of function might especially depend on the dynamic flexibility of neural network.^17^ Furthermore, we expected enhanced prediction performances for dFNC-informed models compared to those that solely employed clinical predictors, such as an initial impairment score, or structural lesion information.

## Methods

### Participants

Fifty-four patients admitted to the University Hospital of Cologne, Department of Neurology, due to acute first-ever ischemic stroke, participated in this study. These patients fulfilled the following inclusion criteria: i) diffusion-weighted imaging (DWI)-verified ischemic stroke, ii) symptom onset less than eight days ago, and iii) 30–99 years of age. Exclusion criteria were: i) hemorrhagic or bi-hemispheric stroke, ii) contraindications to MRI, iii) carotid artery stenosis >50% according to NASCET criteria, or intracranial artery stenosis, iv) cognitive impairment or manifest dementia, v) decreased levels of consciousness, and vi) further neurological or psychiatric diseases. For n = 30 patients, follow-up visits could be scheduled (on average 30 weeks after the ischemic event). All aspects of this study were approved by the ethics committee of the University of Cologne (File No. 11–191), and all patients provided informed written consent following the Declaration of Helsinki. All patients considered here served as replication cohort in Bonkhoff and colleagues 2020.^17^ Furthermore, subsets of the data were previously analyzed with respect to the relationship of acute motor symptoms and connectivity.^8^,^42^ Importantly, these previous analyses considered data from the acute phase only and all analyses presented in this paper are new, hence there is no overlap.

### Motor performance and recovery

Upper limb motor performance was tested twice: at the time of scanning (n=54), i.e., in the acute post-stroke phase and at follow-up six months later. Individual motor performance was captured via the Motricity Index^21^ of the affected upper limb (MI-UL). The MI-UL test assigned scores up to a maximum of 33 points depending on how well movements of (i) shoulder abduction, (ii) elbow flexion, and (iii) pinch grip could be performed (individual scores of the three tests were averaged, 0: No movement, 33: Normal power, c.f. **supplementary table 1**). Patients were categorized in three subgroups depending on the level of motor impairment: (i) patients with no upper limb impairment (MI-UL=33, n=26), (ii) patients with mild impairments (25≤MI-UL≤32, n=16), and (iii) patients with moderate-to-severe upper limb motor impairment (MI-UL≤22, n=12). The cut-off between mildly and moderate-to-severely affected patients was chosen based on the sample distribution of MI-UL scores of patients with impairments and a median score of MI-UL=25.3. As a consequence, patients in the moderate-to-severe group featured an average MI-UL of maximal 22, indicating that at least one movement could not be performed against resistance, equivalent to a clinically relevant motor deficit.

Recovery was quantified as change between follow-up and acute motor impairment (MI- UL_Follow-up_ – MI-UL_acute_). In particular, we defined three subgroups which differed in the amount of experienced motor recovery: (i) no change in motor function (n=16), (ii) more (n=9) and (iii) less pronounced recovery (n=5). The cut-off between more and less pronounced recovery was based on the median amount of recovery in the sample of all patients with an initial impairment (recovery=7.67 MI points). As the difference scores between the different MI categories is on average 6.6, the cut-off value represented a clinically relevant amount of recovery.

### MRI acquisition

Resting-state fMRI data were acquired in the framework of a clinical imaging protocol in a clinical routine on a 1.5T scanner (Philips, Guildford, UK). Patients were asked to lie motionless in the scanner and stay awake. Gradient echo-planar imaging (EPI) parameters were as follows: repetition time (TR)=2,100 ms, echo time (TE)=50 ms, field of view (FOV)=250 mm, 24 axial slices, voxel size: 3.9×3.9×3.9 mm^3^, 183 volumes, acquisition time: **∼**six minutes. We, furthermore, obtained the following structural scans: diffusion-weighted imaging (DWI) images (TR=3,900ms, TE=95ms, FOV=230mm, 22 axial slices, voxel size=1.8×3×6 mm^3^) and T2-weighted MR-images (TR=5,600ms, TE=110ms, FOV=230mm, 22 axial slices, voxel size=0.9×1.1×6.0 mm^3^) for more detailed analyses of lesion topography. Images of patients with right-hemispheric lesions (n=19) were flipped at the midsagittal plane.^8,22^ As a consequence, no conclusions on hemispheric-specific effects can be drawn with respect to motor recovery.

### Structural MRI analysis

Stroke lesion maps were constructed by manually segmenting lesioned tissue on DWI images using MRIcron.^23^ Subsequently, DWI images, as well as corresponding lesion masks, were normalized to standard MNI-space by first co-registering images to an MNI-template and then employing the unified segmentation algorithm after masking infarcted tissue.^24^ Lesion maps of 53 patients passed quality control (in one subject top slices were missing due to an alignment error during DWI volume acquisition). In a subsequent step, we applied principal component analysis (PCA) to reduce the high-dimensional lesioned voxel-space (9,900 voxels lesioned in at least one subject). We performed this PCA-step twice and retained (i) all components that individually explained more than 5% of the variance (*5-component structural lesion data*, 5 components, 64% explained variance in total), and (ii) all components that explained more than 95% of the variance in total (*28-component structural lesion data*, 28 components).^7^

### Resting-state fMRI analysis: Preprocessing

Resting-state fMRI data were preprocessed employing Statistical Parametric Mapping (SPM8; http://www.fil.ion.ucl.ac.uk/spm/) in a Matlab framework (The Mathworks 2012a, Natick, MA, USA). As for one subject, only 182 images were acquired, we shortened all further time courses by one volume to harmonize the scan length across subjects. The first three volumes of each time-series were discarded to allow for blood-oxygenation level dependent (BOLD)- signal saturation.

The 179 remaining images were spatially realigned to the time-series’ mean image and co-registered with the structural image and corresponding lesion mask. Subsequently, all images were spatially normalized to standard MNI-space using the unified segmentation option after masking lesioned brain tissue. In a final step, data were smoothed by a Gaussian kernel with a full-width at half maximum (FWHM) of 8 mm. For each patient’s time series, framewise translation and framewise rotation did not exceed 3 mm and 0.3° maximum, respectively.

### Intrinsic connectivity networks

To define spatially separated intrinsic connectivity networks, components were extracted employing independent component analysis (ICA)^25,26^ on the rsfMRI data of 405 healthy controls (components available for download: http://trendscenter.org/software/).^9,27^ Details on the applied group information guided ICA (“back-reconstruction”) algorithm can be found in Salman *et al*. (2019).^28^

Because of our focus on motor impairments, we centered the analysis on 14 motor network components that can be grouped into three functional domains: Eight cortical sensorimotor components, three subcortical components, and three cerebellar components. Ancillary preprocessing steps comprised time-course de-trending (i.e., accounting for linear, quadratic, and cubic trends in the data), de-spiking using 3Ddespike and application of a fifth-order Butterworth low-pass filter with a high-frequency cutoff of 0.15 Hz. Finally, time-courses were variance normalized.^29^

### Static functional network connectivity

For each subject, we computed ‘classic’ static functional connectivity maps as Fisher’s Z-transformed Pearson’s pairwise correlation of time-courses between all 14 motor networks, resulting in 91 connectivity pairs. Age, sex, mean framewise translation, and rotation were used as independent regressors to correct for demographics and within scanner movement.

### Dynamic functional network connectivity

Subsequently, we estimated dFNC within the framework of the sliding window approach.^9,30,31,10^ As prior studies suggest that sliding window lengths between 30 and 60 seconds allow for a successful dFNC estimation with an optimal signal-to-noise ratio^32^, we opted for a window length of 42 seconds (20 TRs). This step resulted in 159 individual windows that were additionally convolved with a Gaussian of 6.3 seconds (σ=3 TRs). The actual dFNC pairs were obtained from the *l*_1_-regularized precision matrix.^33^ The covariates age, sex, mean framewise translation, and rotation served as regressors-of-no-interest. Finally, dFNC values were normalized by Fisher’s Z-transformation. In the next step, we estimated *connectivity states*, i.e., re-occurring patterns of functional connectivity across time and subject space via k-means clustering^34^ of all patients’ 159 dFNC matrices.^9,10^ We relied on the *l*_1_-distance function given its suitability for high-dimensional data.^35^ In line with previous work,^9,16^ we conducted these clustering processes twice: In the first run, we decided upon the optimal number of clusters *k* (referred to as states). This optimal number *k* was determined based on the elbow criterion, which considers the cluster validity index, computed as the ratio between the within-cluster distance to the between-cluster distance.^9^ In a second clustering run, each of the 159 windows of all 54 patients was assigned to one of *k* connectivity states. By these means, we obtained the following dFNC parameters: *fraction times, dwell times*, and *number of transitions*. Furthermore, we computed the variability of actual dFNC pairs by estimating the standard deviation of pairwise functional connectivity over the 159 windows for each patient.^20^ The larger this value, the more the dynamic connectivity varies over the entire duration of the scan.

### Group differences in dynamic connectivity variability

To further elucidate the nature of the variability in dFNC strength concerning motor impairments, we evaluated differences in the variability of each dynamic connectivity pair between stroke patients with (i) no motor impairment, (ii) mild motor impairment and (iii) and moderate-to-severe motor impairment using a three-level one-way ANOVA (level of significance: *p*<0.05).

In case of significant group differences, we conducted post-hoc t-tests to infer differences between patient groups: no vs. mild impairment, no vs. moderate-to-severe impairment, and mild vs. moderate-to-severe impairment (level of significance: *p*<0.05, FDR-corrected for multiple comparisons). Furthermore, we repeated these analyses steps of three-level one-way ANOVA and post hoc t-tests for the patient sample with follow-up scores.

### Prediction of acute motor impairment and motor recovery

The main aim of the present study was to build robust prediction models of acute individual motor impairment and recovery within the first months based on neuroimaging data acquired in the acute post-stroke phase.

We created two classification scenarios: We aimed at (i) predicting the motor deficits from fMRI data acquired in the acute post-stroke phase and (ii) predicting motor recovery. With respect to the first scenario, we initially sought to determine whether it is possible to predict whether a stroke patient has or doesn’t have a motor deficit using the MI-UL score. In addition, we tested whether it is also possible to even predict the severity of motor impairment, i.e. whether an individual patient had a mild versus a moderate-to-severe impairment of the upper limb. With respect to the second prediction scenario, i.e., predicting motor recovery, the change between the follow-up 6 months post-stroke and the acute MI-UL-score. As we considered three categories of motor recovery (no motor recovery, minor recovery, substantial recovery), we extended previous two-class prediction scenarios to a multi-class prediction one. In an exploratory analysis, we also computed a prediction model to differentiate between patients with minor to no recovery (n=8) and patients with substantial recovery (n=9)

A random forest classifier was used as prediction model.^36^ This meta-estimator machine learning algorithm fits several decision tree classifiers on bootstrapped sub-samples of the dataset and successively averages individual predictions. In this way, it aims to increase prediction accuracy by reducing variance and overfitting. Moreover, random forest classifiers can automatically include non-linear and interaction effects of input variables and handle correlated input variables favorably.^37^ Given our moderate sample size, we avoided an additional nested cross-validation step by adopting hyperparameter settings that were suggested by Olson and colleagues^38^ (n_estimators=500; criterion=“entropy”, max_features=0.25). Olson and colleagues had extracted these settings as the most advantageous ones after running hyperparameter optimizations in 165 biomedical datasets. To obtain a performance estimate for unseen patient data, i.e., the model’s generalization capacity, we conducted 100 randomly initiated five-fold cross-validations, repeatedly developing and evaluating models in 100 × 5 training and test sets. In the multi-class recovery prediction analyses, we employed three-fold cross-validation given the small subgroup sample size.

The reported main performance measure denotes the out-of-sample area under the curve (AUC) and the respective 95%-confidence interval. AUC values range between 0 and 1, and values greater than an AUC=0.5 are considered to be above chance level. Non-overlapping 95%- confidence intervals between models determined significant differences between AUC outcomes. Sensitivity and specificity are given in the supplementary materials. We also examined the feature importance estimated by the random forest classifier to increase the interpretability of the prediction approach. To further elucidate feature importance, we computed Spearman rank correlations between the variables with the highest feature importance and outcomes, i.e., either the acute MI-UL or recovery.

Concerning input features, we examined the predictive capacity of neuroimaging features derived from (i) *structural* data, (ii) *static*, and (iii) *dynamic* functional connectivity data. Structural lesion information was considered either in the 5- or 28-component PCA-reduced form (c.f. **Structural MRI analysis**). Static functional connectivity was entered as 91 network-pair-wise connectivity values. Finally, we constructed two models relying on dFNC. The first dFNC model was based on the dynamic parameters fraction and dwell times, as well as the number of transitions. The second dFNC model leveraged the variability in the dynamic connectivity of all 91 network pairs. Models were built with and without the addition of the acute motricity index when predicting the recovery after stroke.

### Data and code availability

DFNC was computed based on Matlab2019a scripts available in the GIFT toolbox. Further statistical analyses were conducted in a jupyter notebook environment (Python 3.7, https://github.com/AnnaBonkhoff/to_be_added_upon_acceptance)

## Results

### Clinical characteristics

Fifty-four patients participated in this study (mean age: 71.9 (11.8) years, 46% female, days post-stroke: 2.5 (1.5), c.f., **Table 1** for a full list of outcomes and covariates and **Figure 1** for a lesion overlap).

**Table 1.** Demographics, clinical, and MRI characteristics of all 54 stroke patients (mean and standard deviation, if not indicated otherwise). Cortico-spinal tract (CST) affection was computed based on lesion overlap with a CST template provided in the SPM Anatomy Toolbox.^39^

|  | **Acute phase (n=54)** | **Follow-up phase (n=30)** |
| --- | --- | --- |
| **Age** | 71.9 ± 11.8 years | 73.3 ± 10.4 years |
| **Sex (females)** | 46 % | 47 % |
| **Mean Framewise Translation** | 0.22 ± 0.13 mm | 0.24 ± 0.12 mm |
| **Mean Framewise Rotation** | 0.002 ± 0.001 rad | 0.002 ± 0.001 rad |
| **Time since stroke** | 2.5 ± 1.5 days | 210 ± 57 days |
| **Acute: MI UL *affected arm***<br>(median, interquartile range,<br>only patients with an initial<br>deficit) | 25.33 (6.67) (n=28) | 25.33 (4.33) (n=17) |
| **Follow-up: MI UL *affected arm***<br>(median, interquartile range,<br>only patients with an initial<br>deficit) | - | 33.0 (7) (n=17) |
| **Normalized lesion volume**<br>(median, interquartile range) | 2,240 mm<sup>3</sup> (4,768) | 4,048 mm<sup>3</sup> (10,020) |
| **CST overlap** | 1.7 ± 2.0 % | 1.8 ± 2.2 % |

**Figure 1.**
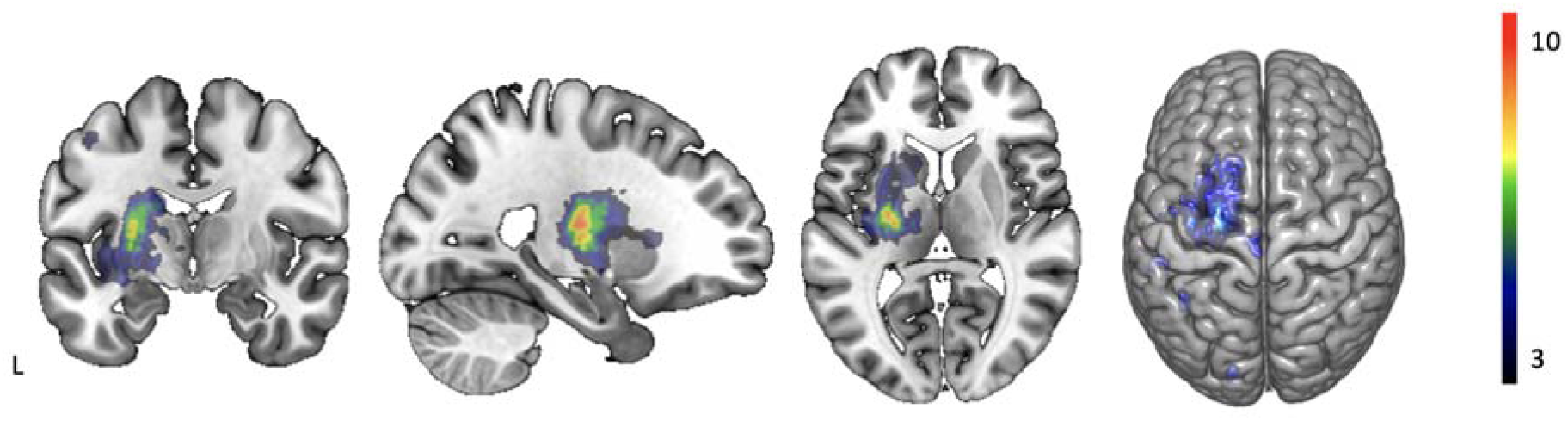
Lesion overlap of all patients. Most lesions were located in the middle cerebral artery territory. The highest lesion load was found subcortically, affecting white matter pathways and the grey matter of basal ganglia.

### Static and dynamic functional network connectivity

After computing time courses and spatial maps of 14 motor components (**Figure 2**), we first estimated static connectivity (**Figure 3A**). Subsequently, we obtained dFNC via the sliding window approach. Hereupon, we identified three discrete, re-occurring connectivity states via k- means clustering, since the cluster validity index suggested three as an optimal cluster number solution (**Figure 3B**).

**Figure 2.**
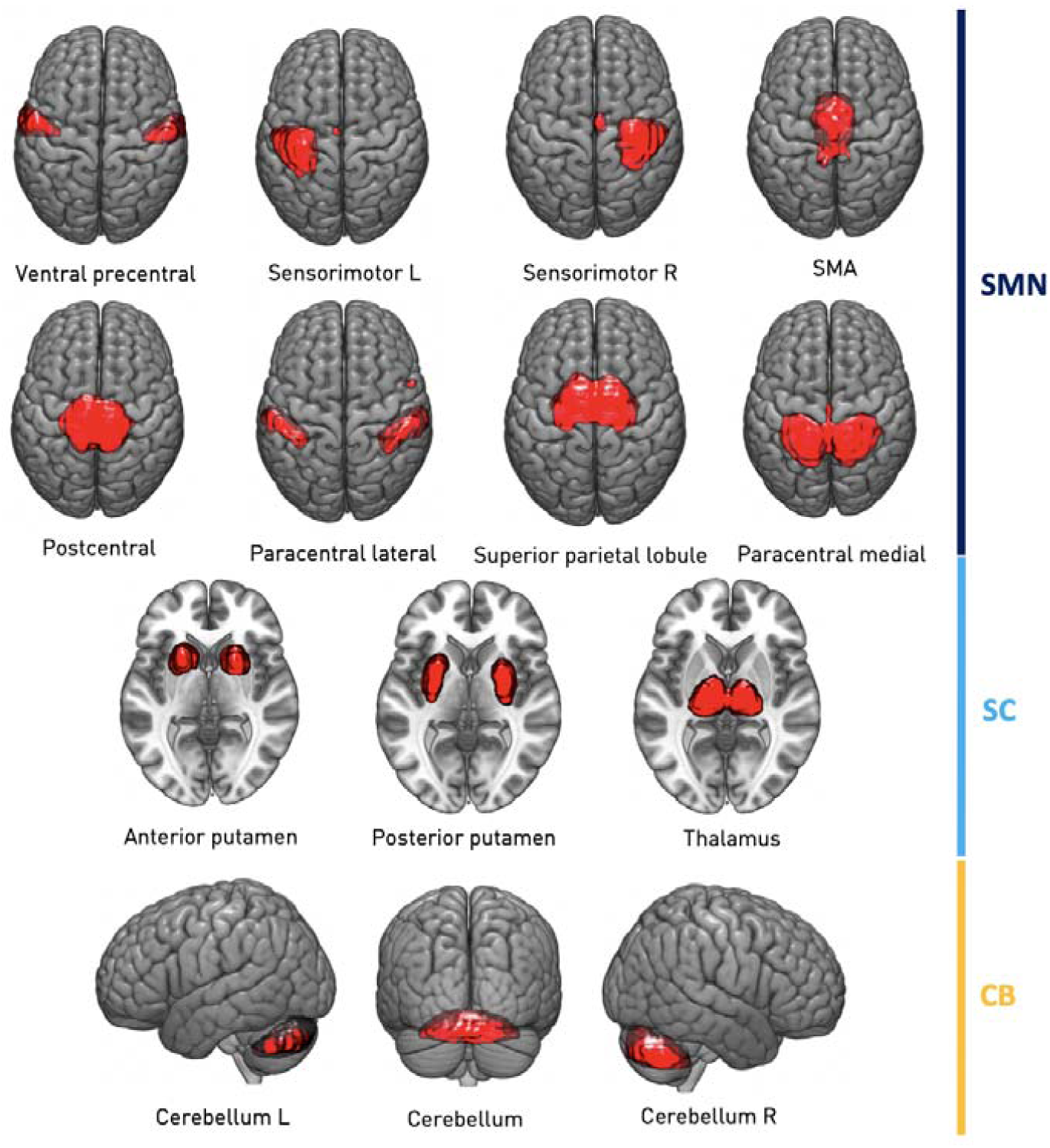
Spatial maps of 14 included intrinsic connectivity networks. Networks were organized in three motor-related functional domains: Sensorimotor (SMN, 8 components, *dark blue*), subcortical (SC, 3 components, *light blue*), and cerebellar (CB, 3 components, *yellow*). Back-reconstruction of networks was based on components extracted in Allen *et al*., 2014. SMA: Supplementary Motor Area. L: Left. R: Right.

**Figure 3.**
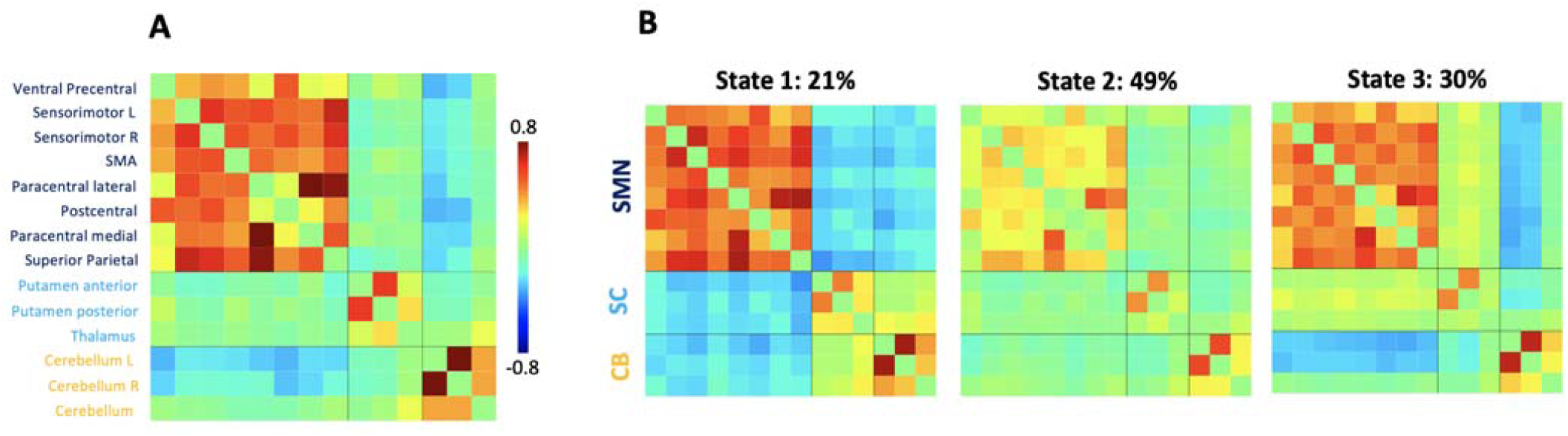
A. Static and B. Dynamic functional network connectivity. **A**. Darker red color implies stronger positive, darker blue stronger negative connectivity. Static functional connectivity was, therefore, characterized by strong positive intra-domain connectivity, neutral connectivity between cortical and subcortical motor networks as well as neutral to negative connectivity between either cortical and subcortical motor networks and cerebellar networks. **B**. Stated percentages above each state correspond to state-specific fraction times across all subjects. State 1, the most seldom state (median dwell time: 10 windows), featured highly positive intra-domain connectivity and highly negative connectivity between the sensorimotor and subcortical as well as cerebellar domains. In contrast, State 2, which emerged most often (median dwell time: 22 windows), was characterized by particularly weak intra-domain connectivity and mostly neutral inter-domain connectivity. Lastly, State 3 presented highly positive intra-domain connectivity, neutral connectivity between the sensorimotor and subcortical, and slightly negative connectivity between sensorimotor and cerebellar domains (median dwell time: 14 windows). We can also notice that visually static functional connectivity resembled State 3 the most. This observation is supported by obtaining the smallest *l*_1_-distance from the static functional connectivity state to all three connectivity states.

### Group differences in dynamic connectivity variability

The variability in dynamic connectivity of eight network pairs differed significantly between the three patient subgroups with different amounts of initial motor impairment (one-way ANOVA: *p*<0.05, **Figure 4A**, left plot). These differences particularly pertained to connections between the cortical sensorimotor networks and the putamen. Mildly affected patients presented with generally lower variability values than both moderately-to-severely and non-affected patients (post hoc t-tests: *p*<0.05, FDR-corrected, **Figure 4B**, outer plots). Moderately-to-severely affected patients presented with even higher dynamic connectivity variability than non-affected patients (post hoc t-tests: *p*<0.05, FDR-corrected, **Figure 4B**, middle plot).

**Figure 4.**
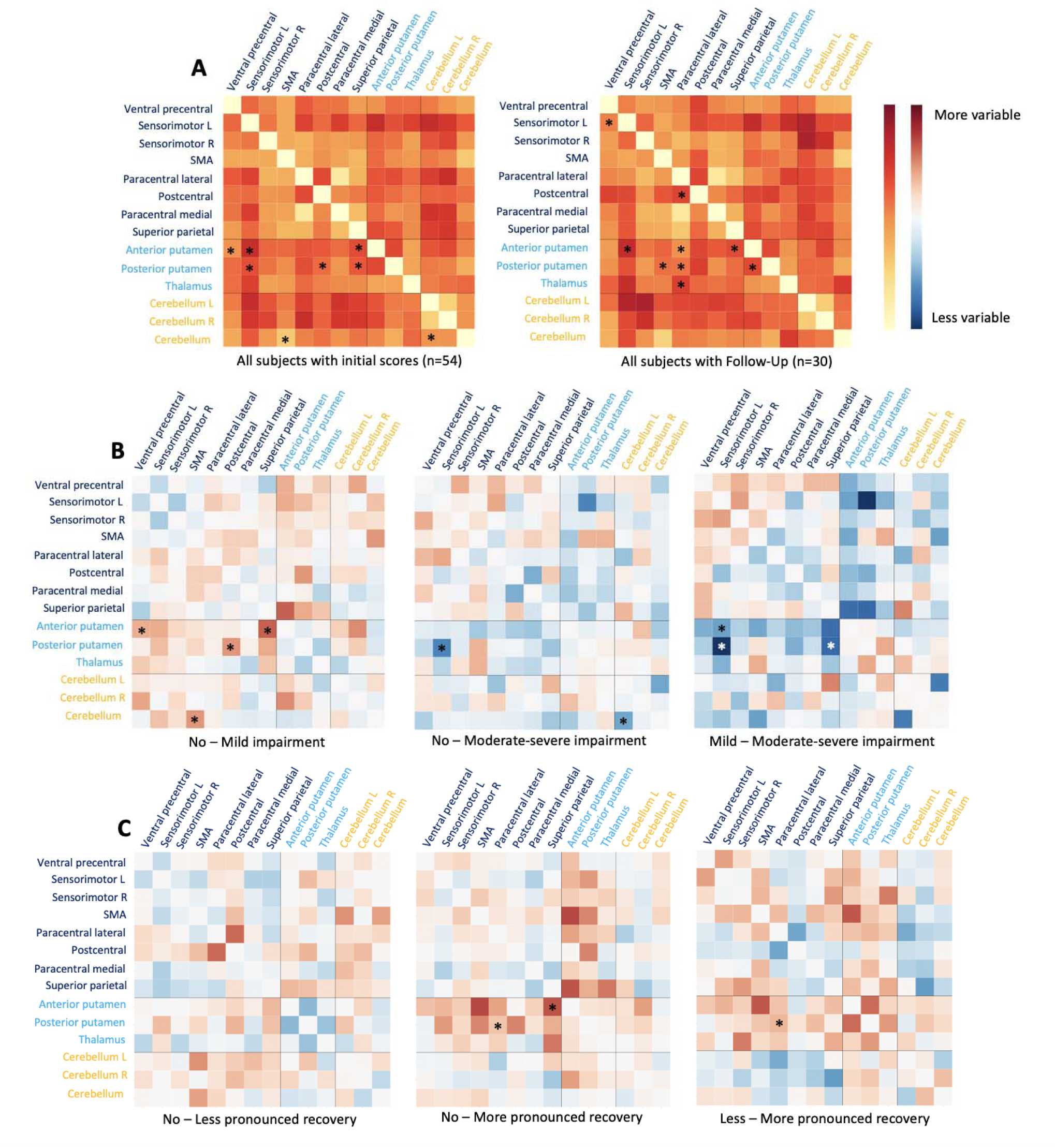
Dynamic functional connectivity strength variability in relation to upper limb motor impairments and recovery post-stroke. A. Mean variability of dynamic connectivity throughout the entire scan session. The left plot visualizes the variability averaged over all 54 patients initially recruited. The right plot considers all 30 patients that were followed up after six months. Darker red color represents higher variability values. Asterisks indicate significant group effects between patients with no, moderate, or severe upper limb impairments or more, less or no recovery, respectively (three-level one-way ANOVA: *p*<0.05). Most of the differences in variability were found for connections between the bilateral putamen and cortical sensorimotor networks. **B. Subtraction maps of mean variability values between each subgroup constellation of initial impairment**. Red color implies higher variability values, blue color lower variability values. Post hoc t-tests revealed some overlapping (“posterior putamen – left sensorimotor cortex”), yet mostly distinct significantly different connectivity pairs between mildely affected patients and non-affected patients, as well as mildly and moderately-to-severely affected patient groups (*p*<0.05, FDR-corrected for multiple comparisons). Mildly affected patients differed from non-affected patients in more dynamic connectivity variability pairs than did moderately-to-severely affected patients (four versus two pairs). **C. Subtraction maps of mean variability values between each subgroup constellation of recoverees**. The asterisk indicated a significant group effect between patients with more pronounced versus those with less pronounced motor recovery post-stroke (t-test: *p*<0.05, FDR-corrected for multiple comparisons).

When contrasting the three patients subgroups with different amounts of *motor recovery* over time, we substantiated nine significantly different dynamic connectivity variability pairs, (one-way ANOVA: *p*<0.05, **Figure 4A**, right plot). Patients in the substantial recovery subgroup were particularly characterized by a lower variability in dynamic connectivity between the supplementary motor area (SMA) and bilateral posterior putamen (post hoc t-tests: *p*<0.05, FDR-corrected, **Figure 4C**, middle and right plots).

### Prediction of acute upper limb impairment

When aiming to predict acute upper limb impairment in the entire sample of 54 stroke subjects, the highest prediction performance yielded the dynamic fraction and dwell times data (out-of-sample AUC±95%-confidence interval(CI): 0.67±0.01). Feature importances indicated that the total time spent in state 3, i.e., the fraction time in a functionally markedly integrated state, was the most relevant variable for predicting the acute impairment status (c.f. **supplemental materials section “Segregation and Integration”**). Dwell times in state 2, another functionally integrated state, were assigned the second highest feature importance. Both features, however, did not correlate with acute motor impairments (fraction time state 3: ρ=0.22, *p*=0.12; dwell time state 2: ρ=0.03, *p*=0.87), which may be indicative of more complex interaction and non-linear effects that the random forest classifier picked up on. The models based on the structural, as well as static functional connectivity data performed second-best (5- component structural: AUC=0.61±0.02, 28-component structural: AUC=0.59±0.01, static connectivity: AUC=0.62±0.02). The model considering the dynamic variability data performed significantly worse than the previous models and achieved an AUC at chance level (**Table 2, Supplementary Table 2**).

**Table 2.** Prediction of acute motor impairment (out-of-sample AUC and 95%-confidence interval). The highest prediction performances per scenario are stated in red and marked with an asterisk

|  | No. of subjects | 5-component structural data | 28-component structural data | Static connectivity data | Faction and dwell times data | Dynamic variability data |
| --- | --- | --- | --- | --- | --- | --- |
| **Acute MI-UL: No-motor impairment vs. motor impairment** | **Structural:** 53<br>(27 vs. 26, 49% without motor symptoms)<br>**fMRI:** 54 | 0.61±0.02 | 0.59±0.01 | 0.62±0.02 | **0.67±0.01\*** | 0.34±0.01 |
|  | (28 vs. 26, 48% without motor symptoms) |  |  |  |  |  |
| **Acute MI-UL: Mild motor impairment vs. moderate-severe motor impairment** | **Structural:** 27 (16 vs. 11, 59% with mild motor impairments)<br>**fMRI:** 28 (16 vs. 12, 57% with mild motor impairments) | 0.22±0.02 | 0.33±0.02 | 0.32±0.01 | 0.34±0.02 | **0.83±0.02\*** |

However, when refining the prediction scenario to the distinction between patients with moderate-to-severe and mild motor impairments (n=28), it was the model based on *dynamic connectivity variabilities* that demonstrated the highest prediction capacity (AUC=0.83±0.02). The amount of motor impairment was especially predicted by the variability of dynamic connectivity between the ipsilesional sensorimotor network and bilateral posterior putamen, as well as ventral precentral cortex and bilateral anterior putamen (**Figure 5)**. Here, more severe motor impairments correlated with higher variability in these networks (ipsilesional sensorimotor network – posterior putamen: ρ=-0.57, *p*=0.001, ventral precentral cortex – anterior putamen: ρ=-0.40, *p*=0.035). None of the other models provided a prediction fidelity above the level of chance for the distinction between impairment levels (**Table 2**).

**Figure 5.**
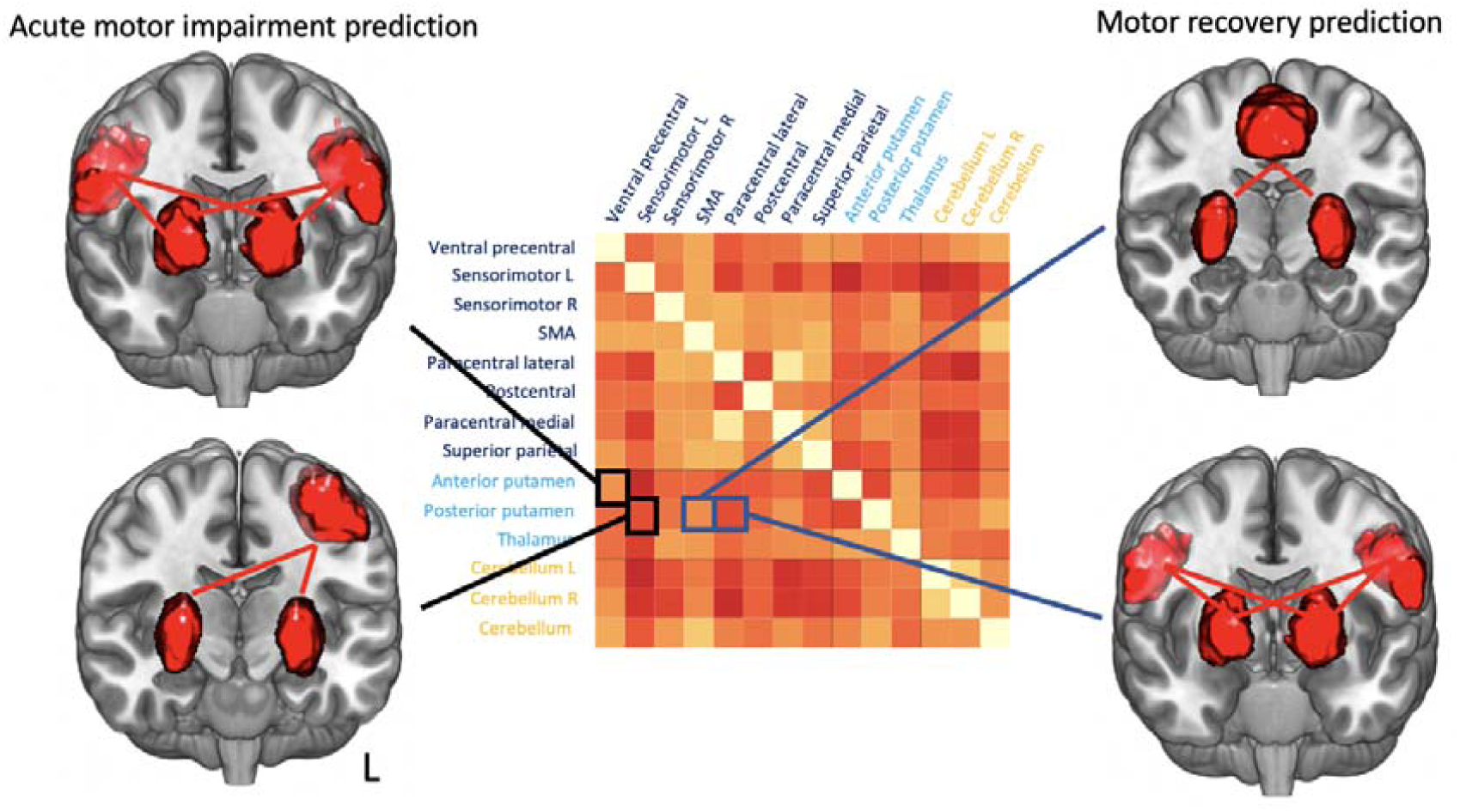
Variability of dynamic connectivity and feature importances. Brain renderings visualize the most predictive dynamic connectivity variability pairs for the prediction of mild versus moderate-severe acute motor impairments (*left*) and the prediction of more or less pronounced recovery in the patients with initial impairment in the first months after stroke (*right*).

In summary, the presence of acute motor impairments was best predicted by the fraction and dwell times-based dynamic connectivity model. In contrast, the variability of individual dynamic connectivity pairs was a powerful predictor of the amount of motor impairment.

### Prediction of motor recovery

We next challenged the capacity of acutely acquired structural and rsfMRI data to predict motor recovery 6 months after stroke (n=30). The acute motricity index was already a strong predictor, when differentiating between more and less pronounced, as well as no changes in motor function in the first six months post-stroke (AUC=0.84±0.01, all results: **Table 3, Supplementary Table 3**). The joint model based on the dynamic fraction and dwell times data and the acute motricity index accomplished a prediction performance of AUC=0.89±0.01. The 95%-confidence intervals of this joint model did not overlap with the 95%-confidence interval of the model using the MI as the sole predictor variable, highlighting significantly improved prediction performance when adding the dynamic connectivity parameters to the behavioral data. The two most important features were the number of transitions between states and fraction time in state 3, i.e., a functionally integrated state. The acute MI-UL score was ranked as the third most important feature only. Both of the most important dynamic connectivity parameters correlated negatively with the recovery, i.e., the more a subject recovered, the shorter their time spent in state 3 was and the fewer they switched between different states (Fraction time state 3: ρ=-0.49, *p*=0.006; Number of state transitions: ρ=-0.39, *p*=0.04).

**Table 3.**
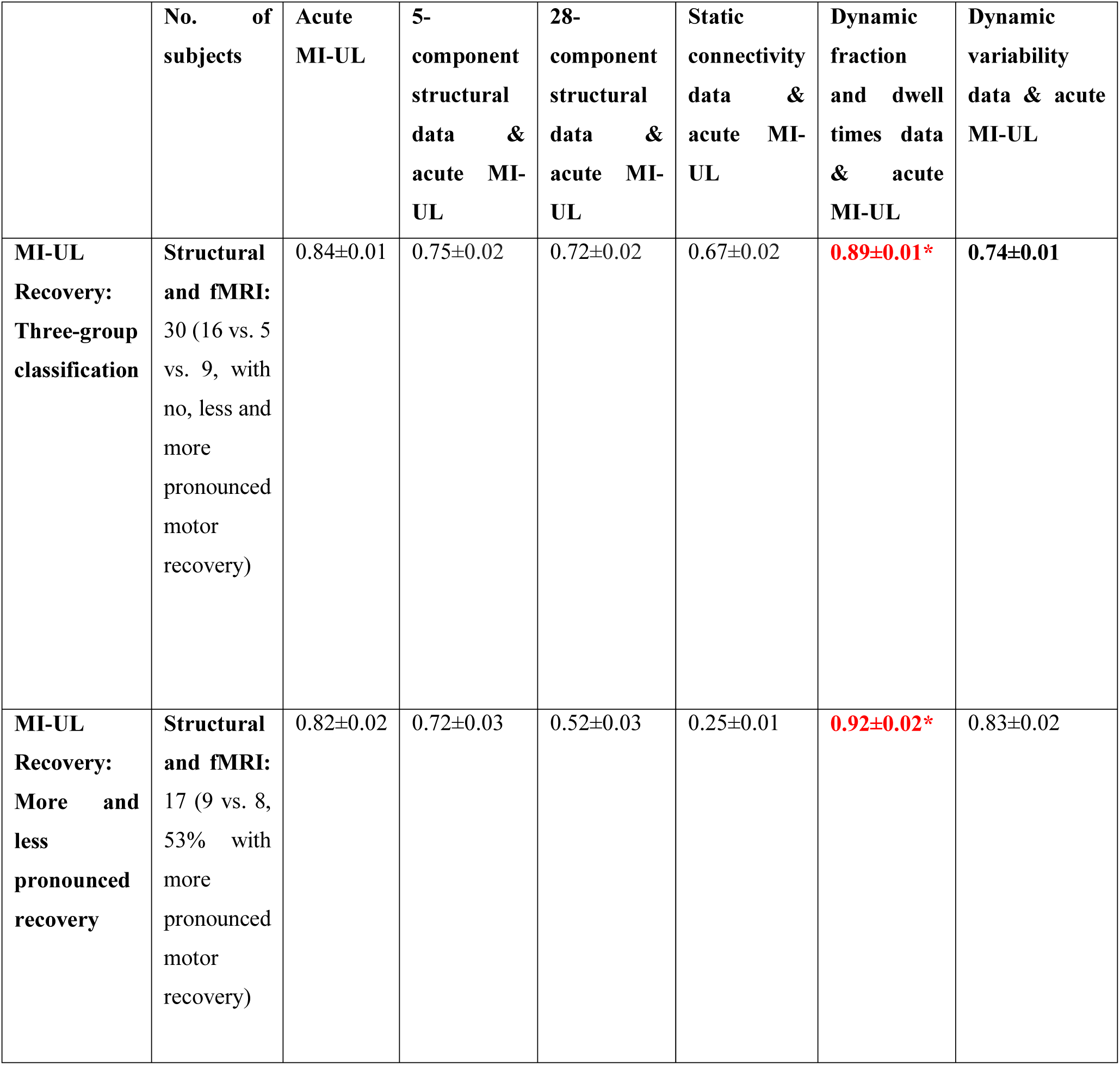
Prediction of upper limb motor recovery based on structural MRI data, static and dynamic connectivity (out-of-sample AUC and 95%-confidence interval). The highest prediction performances per scenario are stated in red and marked with an asterisk.

|  | No. of subjects | Acute MI-UL | 5-component structural data & acute MI-UL | 28-component structural data & acute MI-UL | Static connectivity data & acute MI-UL | Dynamic fraction and dwell times data & acute MI-UL | Dynamic variability data & acute MI-UL |
| --- | --- | --- | --- | --- | --- | --- | --- |
| **MI-UL Recovery: Three-group classification** | **Structural and fMRI:** 30 (16 vs. 5 vs. 9, with no, less and more pronounced motor recovery) | 0.84±0.01 | 0.75±0.02 | 0.72±0.02 | 0.67±0.02 | **0.89±0.01\*** | **0.74±0.01** |
| **MI-UL Recovery: More and less pronounced recovery** | **Structural and fMRI:** 17 (9 vs. 8, 53% with more pronounced motor recovery) | 0.82±0.02 | 0.72±0.03 | 0.52±0.03 | 0.25±0.01 | **0.92±0.02\*** | 0.83±0.02 |

Finally, we tested the prediction performance when only considering patients with an initial impairment. In this scenario, the amount of recovery (no-minor vs. substantial) was best predicted by the dynamic fraction and dwell times data (AUC=0.92±0.02, **Table 3, Supplementary Table 4 & 5**). The most important features were fraction and dwell times in state 3, i.e., a functionally integrated state (Fraction time state 3: ρ=-0.56, *p*=0.02; Dwell time state 3: ρ=-0.61, *p*=0.008). The model relying on the initial MI-score achieved a significantly lower AUC of 0.82 (AUC±95%-CI=0.82±0.02). It was paralleled in performance by the models considering the variability in dynamic connectivity (with the addition of the MI-score: AUC±95%-CI=0.83±0.02 and without the MI-score: AUC±95%-CI=0.82±0.02). The two top ranked features were the variability in dynamic connectivity between SMA and bilateral posterior putamen, as well as lateral paracentral cortex and bilateral posterior putamen. Correlation analyses indicated that subjects showed higher recovery values the smaller variability was in these connections (SMA - putamen: ρ=-0.59, *p*=0.01; lateral paracentral cortex - lateral paracentral cortex: ρ=-0.54, *p*=0.03). While the models relying on 28-component structural and static connectivity data did not exceed a chance-level prediction performance, the model incorporating 5-component structural data had (i) an AUC of 0.72±0.03 with the addition of the MI-score, and (ii) 0.66±0.02 without the MI-score. This finding indicates some relevance of the stroke lesion location with respect to the potential to recover.

In conclusion, we found dFNC parameters obtained in the first days post-stroke such as the state-specific fraction and dwell times as well as the variability of dynamic connectivity to be potent predictors of motor recovery 6 months later. In particular, fraction and dwell times significantly enhanced prediction performance beyond the level of traditional and well-known clinical predictors, such as the initial motor impairment.

## Discussion

We here explored novel predictors of individual acute motor impairment and recovery after stroke. These biomarkers were derived from dFNC analyses of rsfMRI data from 54/30 acute ischemic stroke patients that presented with varying degrees of motor impairment. DFNC analyses are special because they allow for the extraction of moment-to-moment fluctuations in brain connectivity and the definition of re-occurring dynamic connectivity states.^9,10^ Thereby, this approach increases the time resolution of resting-state fMRI signals to seconds and may be particularly capable of capturing stroke-induced short-lasting connectivity alterations and higher network flexibility.^17,19,40^ Fraction times in state 3, i.e., the time spent in a functionally integrated connectivity state, and the variability in dynamic connectivity between putamen and various cortical sensorimotor areas crystallized as particularly promising biomarkers for individualized outcome predictions.

### Functional segregation and integration

Stroke patients with or without acute motor impairments were most accurately differentiated based on the time spent in specific connectivity states. Feature importance highlighted fraction times in state 3, a connectivity state that was characterized by pronounced functional integration^**1**^. Previous studies of dynamic connectivity in independent acute ischemic stroke cohorts have already uncovered links between functional integration and segregation and motor performance: lower motor domain integration in case of severe compared to moderate hand motor impairments^17^ and increased whole-brain segregation (i.e., decreased integration) in case of a high stroke severity.^42^ Interestingly, in healthy subjects with cast-induced motor inactivity of the upper limb, motor network topology also turned into a more segregated state.^43^ We here did not observe a clear correlation between fraction times in integrated state 3 and acute motor impairment across all subjects. Rather, our random forest classifier approach may have enabled us to differentiate between groups by also capturing non-linear, as well as interaction effects between more than one variable.

While we did not find an association between fraction times in state 3 and acute motor impairments, we did, however, identify a high capacity of the fraction and dwell times in state 3 to predict and correlate with future recovery. Patients that did not show a preference for this spatially integrated connectivity state in the first days after stroke recovered more substantially in the upcoming weeks. From a pathomechanistic perspective, the possibly most striking characteristic of state 3 is the pattern of highly positive intra-domain dynamic connectivity in combination with a positive connectivity between cortical and subcortical networks. Hence, the association of lower fraction and dwell times in state 3 with motor recovery indicates that a more segregated, isolated processing within the cortical and subcortical motor domains may be mechanistically critical for a more successful stroke recovery. Interestingly, increased segregation, i.e., decreased integration, has been previously linked to expedited regain of cognitive function in healthy aging and diffusely damaged brains after mild traumatic brain injury.^44^ Our current findings are, therefore, in line with these reports.

### Variability of dynamic connectivity

By considering the variability in dynamic connectivity, we here amend the previously employed toolset to generate insights into the role of motor areas in healthy and pathological motor function. In inference- and prediction-focused analyses, we were able to link mild motor impairment as well as a more pronounced recovery to lower, i.e., more stable, dynamic connectivity variability values in select cortical – subcortical connections. The investigation of dynamic connectivity variability profiles was previously motivated by Allen and colleagues,^9^ especially as they represented inaccessible information content when conducting static connectivity analyses. The authors described distinct “zones of instability”, i.e., regions with more variable dynamic connectivity in healthy volunteers: The most variable regions were located in the lateral parietal and occipital cortex. Comparable analyses of dFNC variability in neurological patients with temporal lobe epilepsy inferred the instability of precuneus dynamic connectivity as a signature of the disease.^45^ In yet another study, the dFNC variability provided powerful information for the differentiation between Alzheimer’s patients and healthy controls.^20^ In the present study, it was the variability of the dynamic connectivity between the ipsilesional sensorimotor cortex and bilateral posterior putamen that predicted a more versus less initial impairment especially well. Statistical group comparisons moreover suggested that, both moderately-to-severely affected patients, as well as those without any impairments had a higher ipsilesional sensorimotor-putamen dFNC variability than mildly affected patients. Both of these brain regions are well known to be implicated in the emergence of acute motor impairments and recovery post-stroke: Lesion symptom mapping studies repeatedly reported ischemic lesions in the putamen underlying upper limb impairment.^46,47^ Furthermore, the link between lower (static) resting-state connectivity between ipsi- and contralesional motor areas and higher motor impairment is one of the most prominently featured findings in stroke neuroimaging research.^8,48^ The connectivity between the posterior ipsilesional sensorimotor cortex and putamen, let alone their dFNC variability, however, requires further exploration to determine the biological meaning.

In case of predicting motor recovery, the variability of the dynamic connectivity strengths between the supplementary motor areas (SMA) and the bilateral posterior parts of the putamen was the most predictive feature. This SMA-putamen variability was also found to be significantly lower in case of more pronounced recovery compared to less pronounced and no change in motor function. The SMA has been consistently identified as critical region for physiological motor function as well as for stroke recovery in previous studies.^49,50^ In particular, longitudinal motor-task-based functional imaging studies demonstrated a recovery-related excess in activation in these regions early after stroke^49^ and decreases in activation in sub-acute and chronic stages after stroke.^50^ In addition, dynamic causal modelling (DCM) analyses that extracted links between increases in SMA-M1 coupling and motor improvements suggested a supportive role of SMA for motor recovery post-stroke.^51^ Lastly, diffusion-tensor imaging in healthy adults indicates structural connections between SMA and especially posterior parts of the putamen, highlighting the role of this connection for motor function.^52^ In line with this conclusion, the SMA-putamen connectivity was found to be disturbed in patients suffering from motor impairments due to Parkinson’s disease.^53,54^

In summary, previous results on dFNC variability and ours combined suggest that these dFNC parameters represent biologically meaningful fingerprints of neurological diseases. In the case of ischemic stroke, this variability might particularly well capture the effects of brain injury and early plasticity mechanism in the first few days post-stroke.

### Limitation and future directions

A sample of 54 stroke subjects is arguable still extendible if intended for the construction of outcome prediction models, especially as the sample size was decreased further in ancillary analyses. However, the present dataset is among the largest acute stroke rsfMRI datasets currently available and rendered particularly unique by its comprehensive longitudinal motor assessment and acquisition during the clinical routine, i.e., when the patients received their diagnostic MR scans. The latter circumstance enabled a very early time point of data acquisition, on average 2.5 days after stroke. This underlines the general feasibility of implementing the approach presented here with respect to clinical translation. Due to the task-free nature of rsfMRI, it was also feasible to recruit stroke patients that are typically excluded from fMRI task studies, i.e., patients with severe motor impairment. Lastly, we have focused on classification scenarios, i.e., our prediction models discriminated patients with or without motor impairments. It could thus be a valuable next step to predict outcomes on continuous scales, i.e., predict the precise amount of impairment per patient.

## Conclusions

Here, we demonstrated the feasibility of predicting acute motor impairment and recovery after stroke based on dynamic functional connectivity. Individual patients’ preferences for a highly integrated connectivity state were particularly pivotal in discriminating between the presence or absence of acute motor impairment and recovery over time. We additionally uncovered significantly different profiles in the variability of dynamic connectivity between patient groups with varying degrees of motor impairment. These differences especially related to links between the putamen and cortical motor networks, some of which also emerged as reliable predictors of motor impairment and recovery. In conclusion, our study highlights the value of dynamic connectivity-derived information to gain insights into the phenotypes of acute ischemic brain injury and recovery after stroke.

## Data Availability

DFNC was computed based on Matlab2019a scripts available in the GIFT toolbox. Further statistical analyses were conducted in a jupyter notebook environment (Python 3.7, https://github.com/AnnaBonkhoff/to_be_ADDED_afterpublication).

## Acknowledgments

We are grateful to our colleagues at the Department of Neurology, University Hospital Cologne & Medical Faculty, University of Cologne, for valuable support and discussions. Furthermore, we are grateful to our research participants without whom this work would not have been possible.

## Author contributions

C.G., A.K.R., A.K.B.: Conception and design of study; C.G., V.C., V.M.V., H.G., F.A.E., A.K.R., A.K.B.: Acquisition and analysis of data; C.G., N.S.R., V.C., G.R.F., L.J.V., C.T., L.H., A.K.R., A.K.B.: Drafting of significant portions of the manuscripts and figures; all authors: editing and approving the text.

## Abbreviations

AUC: Area under curve
dFNC: dynamic functional network connectivity
FDR: false discovery rate
MRI: magnetic resonance imaging
MI-UL: motricity index of the upper limb
rsfMRI: resting-state functional magnetic resonance imaging

## Supplementary materials

**Supplementary Table 1A.** Motricity index: Shoulder abduction and elbow flexion.

|  |  |
| --- | --- |
| **0** | No movement |
| **9** | Palpable contraction of the muscle, but no movement |
| **14** | Visible movement, but not full range and not against gravity |
| **19** | Full range of movement against gravity, but not resistance |
| **25** | Full movement against gravity, but weaker than on the other side |
| **33** | Normal power |

**Supplementary Table 1B.** Motricity index: Pinch grip. Please note that we took the average of all three scores to arrive at our maximum score of 33 for the entire MI-UL.

|  |  |
| --- | --- |
| **0** | No movement |
| **11** | Beginning of prehension |
| **19** | Able to grip cube, but not hold it against gravity, examiner may need to lift the wrist |
| **22** | Able to grip and hold the cube against gravity |
| **26** | Able to grip and hold the cube against a weak pull, but weaker than on the other side |
| **33** | Normal power |

### Segregation and Integration

We computed domain-wide segregation scores for each of the three derived dynamic connectivity states utilizing the formula:^55^

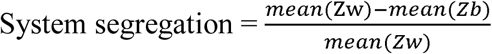

with Mean(Z_w_): average of all within-domain correlations (measured as Fisher’s Z-transformed *r*) and mean(Z_b_): average of all between-domain correlations. Negative correlation values to zero in accordance with Chan *et al*. (2014).

State 1 was found to be the most segregated state (State 1: 0.92, averaged over all subjects), while state 2 and 3 were similarly integrated (State 2: 0.81, State 3: 0.82; one-way ANOVA comparing domain-wide segregation of all three states: F-value=21.8, *p*<0.001; post hoc t-tests: State 1 – State 2: T-value=6.26, *p*<0.01, State 1 – State 3: T-value=6.39, *p*<0.001, State 2 – State 3: T-value=-0.06, *p*=0.95).

**Supplementary Table 2.** Prediction of acute motor impairment (test-set sensitivity and specificity as well as 95%-confidence intervals).

|  | No. of subjects | 5-component structural data | 28-component structural data | Static connectivity data | Dynamic fraction and dwell times data | Dynamic variability data |
| --- | --- | --- | --- | --- | --- | --- |
| **Acute MI-UL: No-motor impairment vs. motor impairment** | **Structural:**<br>53<br>(27 vs. 26, 49% without motor symptoms) | Sens=0.54±0.0<br>2<br>Spec=0.61±0.0<br>2 | Sens=0.56±0.0<br>2<br>Spec=0.60±0.0<br>2 | Sens=0.57±0.0<br>2<br>Spec=0.64±0.0<br>2 | Sens=0.64±0.0<br>2<br>Spec=0.57±0.0<br>2 | Sens=0.33±0.0<br>2<br>Spec=0.44±0.0<br>2 |
|  | **fMRI:** 54<br>(28 vs. 26,<br>48%<br>without<br>motor<br>symptoms) |  |  |  |  |  |
| **Acute MI-**<br>**UL: Mild**<br>**motor**<br>**impairme**<br>**nt vs.**<br>**moderate-**<br>**severe**<br>**motor**<br>**impairme**<br>**nt** | **Structural:**<br>27 (16 vs.<br>11, 59%<br>with mild<br>motor<br>impairment<br>s)<br>**fMRI:** 28<br>(16 vs. 12,<br>57% with<br>mild motor<br>impairment<br>s) | Sens=0.53±0.0<br>3<br>Spec=0.12±0.0<br>2 | Sens=0.63±0.0<br>3<br>Spec=0.22±0.0<br>2 | Sens=1.00±0.0<br>0<br>Spec=0.50±0.0<br>0 | Sens=0.66±0.0<br>2<br>Spec=0.20±0.0<br>2 | Sens=0.86±0.0<br>2<br>Spec=0.59±0.0<br>3 |

**Supplementary table 3.** Prediction of motor recovery based on structural MRI data and static as well as dynamic connectivity alone (test-set AUC, 95%-confidence intervals).

|  | No. of<br>subjects | 5-component<br>structural data | 28-component<br>structural data | Static<br>connectivity<br>data | Dynamic<br>fraction and<br>dwell times<br>data | Dynamic<br>variability data |
| --- | --- | --- | --- | --- | --- | --- |
| **MI-UL**<br>**Recovery:**<br>**Three-group**<br>**classification** | **Structural**<br>**and fMRI:**<br>30 (16 vs. 5<br>vs. 9, with<br>no, less and<br>more<br>pronounced motor recovery) | AUC=0.63±0.02 | AUC=0.54±0.01 | AUC=0.52±0.01 | AUC=0.77±0.01 | AUC=0.64±0.01 |

**Supplementary Table 4.**
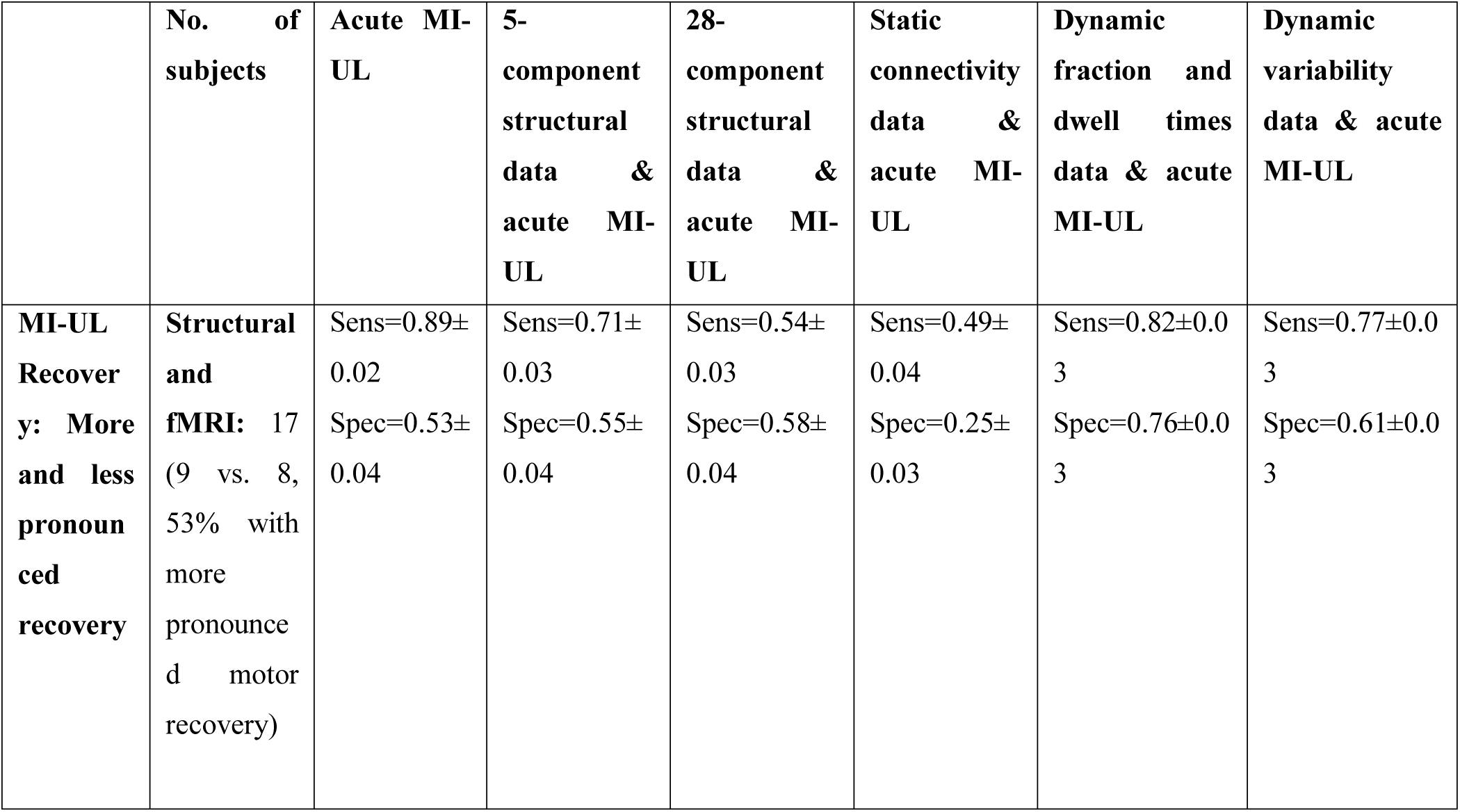
Prediction of motor recovery in the sample of all patients with initial motor impairment (test-set sensitivity and specificity as well as 95%-confidence intervals).

**Supplementary Table 5.** Prediction of motor recovery in the sample of all patients with initial motor impairment based on structural MRI data and static as well as dynamic connectivity alone (test-set AUC, sensitivity, specificity and respective 95%-confidence intervals).

|  | No. of subjects | 5-component structural data | 28-component structural data | Static connectivity data | Fraction and dwell times data | Dynamic variability data |
| --- | --- | --- | --- | --- | --- | --- |
| **MI-UL** | **Structural** | AUC=0.66± | AUC=0.52± | AUC=0.22± | AUC=0.91±0. | AUC=0.82±0. |
| **Recovery: More and less pronounced recovery** | **and fMRI:** 17 (9 vs. 8, 53% with more pronounced motor recovery) | 0.03<br>Sens=0.65±0.03<br>Spec=0.45±0.04 | 0.03<br>Sens=0.56±0.03<br>Spec=0.47±0.04 | 0.03<br>Sens=0.40±0.04<br>Spec=0.29±0.04 | 02<br>Sens=0.79±0.03<br>Spec=0.76±0.03 | 02<br>Sens=0.76±0.03<br>Spec=0.64±0.04 |

## References

1. Benjamin, E. J. et al. Heart disease and stroke statistics-2017 update: a report from the American Heart Association. Circulation 135, e146–e603 (2017).

2. Hay, S. I. et al. Global, regional, and national disability-adjusted life-years (DALYs) for 333 diseases and injuries and healthy life expectancy (HALE) for 195 countries and territories, 1990–2016: a systematic analysis for the Global Burden of Disease Study 2016. The Lancet 390, 1260–1344 (2017).

3. Prabhakaran, S. et al. Inter-individual Variability in the Capacity for Motor Recovery After Ischemic Stroke. Neurorehabilitation and Neural Repair 22, 64–71 (2008).

4. Kundert, R., Goldsmith, J., Veerbeek, J. M., Krakauer, J. W. & Luft, A. R. What the Proportional Recovery Rule Is (and Is Not): Methodological and Statistical Considerations. Neurorehabilitation and neural repair 1545968319872996 (2019).

5. Bonkhoff, A. K. et al. Bringing proportional recovery into proportion: Bayesian modelling of post-stroke motor impairment. Brain (2020) doi:10.1093/brain/awaa146.

6. Rehme, A. K. et al. Individual prediction of chronic motor outcome in the acute post□stroke stage: Behavioral parameters versus functional imaging. Human brain mapping 36, 4553–4565 (2015).

7. Siegel, J. S. et al. Disruptions of network connectivity predict impairment in multiple behavioral domains after stroke. Proceedings of the National Academy of Sciences 113, E4367–E4376 (2016).

8. Rehme, A. K. et al. Identifying neuroimaging markers of motor disability in acute stroke by machine learning techniques. Cerebral cortex 25, 3046–3056 (2015).

9. Allen, E. A. et al. Tracking Whole-Brain Connectivity Dynamics in the Resting State. Cerebral Cortex 24, 663–676 (2014).

10. Calhoun, V. D., Miller, R., Pearlson, G. & Adalı, T. The Chronnectome: Time-Varying Connectivity Networks as the Next Frontier in fMRI Data Discovery. Neuron 84, 262–274 (2014).

11. Vidaurre, D., Arenas, A. L., Smith, S. M. & Woolrich, M. W. Behavioural relevance of spontaneous, transient brain network interactions in fMRI. bioRxiv 779736 (2019).

12. Lurie, D. J. et al. On the nature of resting fMRI and time-varying functional connectivity. Advance online publication. Retrieved December 24, 2018 (2018).

13. Liu, F. et al. Dynamic functional network connectivity in idiopathic generalized epilepsy with generalized tonic-clonic seizure: Dynamic FNC in IGE-GTCS. Human Brain Mapping 38, 957–973 (2017).

14. Tu, Y. et al. Abnormal thalamocortical network dynamics in migraine. Neurology 92, e2706–e2716 (2019).

15. Kim, J. et al. Abnormal intrinsic brain functional network dynamics in Parkinson’s disease. Brain 140, 2955–2967 (2017).

16. Espinoza, F. A. et al. Whole-Brain Connectivity in a Large Study of Huntington’s Disease Gene Mutation Carriers and Healthy Controls. Brain Connectivity 8, 166–178 (2018).

17. Bonkhoff, A. K. et al. Acute ischaemic stroke alters the brain’s preference for distinct dynamic connectivity states. Brain (2020) doi:10.1093/brain/awaa101.

18. Rashid, B. et al. Classification of schizophrenia and bipolar patients using static and dynamic resting-state fMRI brain connectivity. NeuroImage 134, 645–657 (2016).

19. Vergara, V. M., Mayer, A. R., Kiehl, K. A. & Calhoun, V. D. Dynamic functional network connectivity discriminates mild traumatic brain injury through machine learning. NeuroImage: Clinical 19, 30–37 (2018).

20. de Vos, F. et al. A comprehensive analysis of resting state fMRI measures to classify individual patients with Alzheimer’s disease. NeuroImage 167, 62–72 (2018).

21. Demeurisse, G., Demol, O. & Robaye, E. Motor evaluation in vascular hemiplegia. European neurology 19, 382–389 (1980).

22. Volz, L. J. et al. Shaping early reorganization of neural networks promotes motor function after stroke. Cerebral cortex 26, 2882–2894 (2016).

23. Rorden, C. & Brett, M. Stereotaxic display of brain lesions. Behavioural neurology 12, 191–200 (2000).

24. Ashburner, J. & Friston, K. J. Unified segmentation. Neuroimage 26, 839–851 (2005).

25. Lin, Q.-H., Liu, J., Zheng, Y.-R., Liang, H. & Calhoun, V. D. Semiblind spatial ICA of fMRI using spatial constraints. Human brain mapping 31, 1076–1088 (2010).

26. Du, Y. & Fan, Y. Group information guided ICA for fMRI data analysis. Neuroimage 69, 157–197 (2013).

27. Calhoun, V. D., Adali, T., Pearlson, G. D. & Pekar, J. J. A method for making group inferences from functional MRI data using independent component analysis. Human brain mapping 14, 140–151 (2001).

28. Salman, M. S. et al. Group ICA for identifying biomarkers in schizophrenia:’Adaptive’networks via spatially constrained ICA show more sensitivity to group differences than spatio-temporal regression. NeuroImage: Clinical 22, 101747 (2019).

29. Rachakonda, S., Egolf, E., Correa, N. & Calhoun, V. Group ICA of fMRI toolbox (GIFT) manual. Dostupnéz http://www.nitrc.org/docman/view.php/55/295/v1.3d_GIFTManualpdf[cit.2011-11-5] (2007).

30. Damaraju, E. et al. Dynamic functional connectivity analysis reveals transient states of dysconnectivity in schizophrenia. NeuroImage: Clinical 5, 298–308 (2014).

31. Sakoğlu, Ü. et al. A method for evaluating dynamic functional network connectivity and task-modulation: application to schizophrenia. Magnetic Resonance Materials in Physics, Biology and Medicine 23, 351–366 (2010).

32. Preti, M. G., Bolton, T. A. & Van De Ville, D. The dynamic functional connectome: State-of-the-art and perspectives. Neuroimage 160, 41–54 (2017).

33. Friedman, J., Hastie, T. & Tibshirani, R. Sparse inverse covariance estimation with the graphical lasso. Biostatistics 9, 432–441 (2008).

34. Lloyd, S. Least squares quantization in PCM. IEEE transactions on information theory 28, 129–137 (1982).

35. Aggarwal, C. C., Hinneburg, A. & Keim, D. A. On the surprising behavior of distance metrics in high dimensional space. in International conference on database theory 420–434 (Springer, 2001).

36. Breiman, L. Random forests. Machine learning 45, 5–32 (2001).

37. James, G., Witten, D., Hastie, T. & Tibshirani, R. An introduction to statistical learning. vol. 112 (Springer, 2013).

38. Olson, R. S., Cava, W. L., Zairah Mustahsan, Varik, A. & Moore, J. H. Data-driven advice for applying machine learning to bioinformatics problems. Pacific Symposium on Biocomputing. Pacific Symposium on Biocomputing 23, 192–203 (2018).

39. Eickhoff, S. B. et al. A new SPM toolbox for combining probabilistic cytoarchitectonic maps and functional imaging data. Neuroimage 25, 1325–1335 (2005).

40. Horn, H. J. van der et al. Functional outcome is tied to dynamic brain states after mild to moderate traumatic brain injury. Human Brain Mapping 41, 617–631 (2020).

41. Friston, K. J. Functional and Effective Connectivity: A Review. Brain Connectivity 1, 13–36 (2011).

42. Bonkhoff, A. K. et al. Dynamic functional connectivity analysis reveals transiently increased segregation in patients with severe stroke. medRxiv (2020).

43. Newbold, D. J. et al. Plasticity and Spontaneous Activity Pulses in Disused Human Brain Circuits. Neuron (2020) doi:10.1016/j.neuron.2020.05.007.

44. Gallen, C. L. & D’Esposito, M. Brain modularity: A biomarker of intervention-related plasticity. Trends in cognitive sciences (2019).

45. Robinson, L. F. et al. The temporal instability of resting state network connectivity in intractable epilepsy. Human brain mapping 38, 528–540 (2017).

46. Sperber, C., Rennig, J. & Karnath, H.-O. Neural correlates of imaging biomarkers for primary motor deficits after stroke – more than just the primary motor system? http://biorxiv.org/lookup/doi/10.1101/2020.07.20.212175 (2020) xdoi:10.1101/2020.07.20.212175.

47. Findlater, S. E. et al. Comparing CST Lesion Metrics as Biomarkers for Recovery of Motor and Proprioceptive Impairments After Stroke. Neurorehabil Neural Repair 33, 848–861 (2019).

48. Carter, A. R. et al. Resting interhemispheric functional magnetic resonance imaging connectivity predicts performance after stroke. Annals of neurology 67, 365–375 (2010).

49. Rehme, A. K., Fink, G. R., von Cramon, D. Y. & Grefkes, C. The role of the contralesional motor cortex for motor recovery in the early days after stroke assessed with longitudinal FMRI. Cerebral cortex 21, 756–768 (2011).

50. Ward, N. S. Neural correlates of motor recovery after stroke: a longitudinal fMRI study. Brain 126, 2476–2496 (2003).

51. Rehme, A. K., Eickhoff, S. B., Wang, L. E., Fink, G. R. & Grefkes, C. Dynamic causal modeling of cortical activity from the acute to the chronic stage after stroke. NeuroImage 55, 1147–1158 (2011).

52. Lehéricy, S. et al. Diffusion tensor fiber tracking shows distinct corticostriatal circuits in humans. Annals of Neurology 55, 522–529 (2004).

53. Wu, T. et al. Effective connectivity of brain networks during self-initiated movement in Parkinson’s disease. NeuroImage 55, 204–215 (2011).

54. Yu, R., Liu, B., Wang, L., Chen, J. & Liu, X. Enhanced Functional Connectivity between Putamen and Supplementary Motor Area in Parkinson’s Disease Patients. PLoS One 8, (2013).

55. Chan, M. Y., Park, D. C., Savalia, N. K., Petersen, S. E. & Wig, G. S. Decreased segregation of brain systems across the healthy adult lifespan. Proceedings of the National Academy of Sciences 111, E4997–E5006 (2014).

